# The Dermatology Life Quality Index is a useful patient reported outcome measure in individuals with severe erythema nodosum leprosum: a post-hoc analysis of the Methotrexate and Prednisolone study - MaPs in ENL

**DOI:** 10.64898/2026.05.21.26353785

**Authors:** Barbara de Barros, Neeta Maximus, Farha Sultana, Bishwanath Acharya, Vivek V. Pai, Anju Wakade, Bhagyashree Bhame, Abdulnaser Hamza, Alemtsehay Getachew, Medhi Denisa Alinda, M. Yulianto Listiawan, Shimelis N Doni, Deanna A. Hagge, Indra Napit, Mahesh Shah, Joydeepa Darlong, Peter Nicholls, Bernd Genser, Saba M. Lambert, Diana N. J. Lockwood, Stephen L. Walker, the Erythema Nodosum Leprosum International STudy Group

## Abstract

**BACKGROUND:** Erythema nodosum leprosum (ENL) is a severe inflammatory complication of leprosy associated with disability, morbidity and mortality. Impairment of health-related quality of life (HRQoL) in ENL has been reported using the Dermatology Life Quality Index (DLQI) and the 36-Item Short Form Health Survey (SF-36), the latter validated in people affected by leprosy. Understanding the correlation between these measures is important to determine whether the shorter dermatology-specific DLQI provides a valid and practical measure of HRQoL in ENL.

**OBJECTIVES:** To examine the relationship between DLQI and SF-36 scores in individuals with ENL using data from the Methotrexate and Prednisolone study in ENL (MaPs in ENL).

**METHODS:** A post-hoc analysis of prospectively collected HRQoL data from the trial sites in India, Indonesia, and Nepal of the MaPs in ENL multicentre randomised clinical trial was performed. HRQoL was assessed using the DLQI and SF-36 at enrolment and at weeks 24, 48 and 60. Associations between DLQI and SF-36 physical (PCS) and mental (MCS) component summary scores were evaluated using correlation analyses and multivariable linear regression at enrolment, and linear mixed-effects models during follow-up adjusted for age, sex, recruiting centre and enrolment SF-36 scores.

**RESULTS:** A total of 383 paired HRQoL assessments from 129 participants were analysed. At enrolment, HRQoL impairment was substantial (median DLQI 19, IQR 15–21; mean PCS 30.3 ±7.3; mean MCS 33.3 ±8.4). DLQI scores improved markedly during follow-up. Across all timepoints, DLQI was strongly inversely correlated with PCS (r = −0.848) and MCS (r = −0.806) (both p<0.001). In adjusted analyses, higher DLQI scores were consistently associated with lower PCS and MCS. At enrolment, each 1-point increase in DLQI was associated with a 0.66-point reduction in PCS and a 0.51-point reduction in MCS (both p<0.001). These associations remained strong during follow-up, with no evidence that they varied over time.

**CONCLUSIONS:** DLQI scores were strongly and consistently associated with SF-36 physical and mental health scores. These findings support the use of the DLQI as a practical patient reported outcome measure to assess the HRQoL associated with ENL and its change following treatment.

**What is already known about this topic?:**

- ENL is associated with substantial morbidity and impaired quality of life.
- Dermatology-specific and generic HRQoL instruments have been used in leprosy.
- Comparative data between DLQI and SF-36 in ENL are scarce and limited.

**What does this study add?:**

- The DLQI strongly inversely correlated with with the SF-36 measures in ENL.
- Dermatology-specific impairment reflects broader physical and mental health burden in ENL.
- The DLQI may be a practical outcome measure for ENL clinical studies and routine clinical assessment.

## INTRODUCTION

Leprosy is a stigmatising, chronic granulomatous disease caused by *Mycobacterium leprae* and *Mycobacterium lepromatosis* that primarily affects the skin and peripheral nerves. Although curable with multidrug therapy, leprosy remains a public health problem in many endemic countries because transmission persists, new cases continue to occur, and delayed diagnosis still results in preventable disability. In 2024, 172,717 new cases were reported globally, with 23 World Health Organization (WHO) priority countries accounting for most of the burden (1–3). Clinical manifestations occur along an immunological spectrum determined by the host immune response, ranging from tuberculoid to lepromatous disease. The cardinal signs of leprosy include hypopigmented or erythematous skin lesions with definite sensory loss, thickened peripheral nerves with associated sensory or motor impairment, and the demonstration of acid-fast bacilli in slit-skin smears (2). Skin lesions are the commonest presenting feature of leprosy(1,2,4), whilst peripheral neuropathy is also frequent and may already be evident at diagnosis. In one Ethiopian cohort of multibacillary leprosy, 84% of new cases had at least one thickened peripheral nerve and up to 55% had nerve function impairment at presentation (5).

Disability remains a major consequence of leprosy and arises mainly from peripheral nerve damage caused by neural mycobacterial infiltration and by host immune-mediated inflammation directed at mycobacterial antigen (6–8). Sensory loss, muscle weakness and autonomic dysfunction may result in trauma, ulceration and deformity, particularly affecting the hands, feet and eyes. WHO classifies disability in leprosy using a three-grade system (0-2): grade 0 indicates no anaesthesia or visible deformity; grade 1 indicates loss of sensation without visible deformity; and grade 2 indicates visible deformity or damage (e.g. ulcers, claw hand, lagophthalmos) (9). Grade 2 disability at diagnosis is widely used as an indicator of delayed case detection (10).

Stigma is a major determinant of health and quality of life in leprosy (1,2). Visible skin lesions, deformity, treatment-related skin discolouration and unfounded fear of contagion may contribute to enacted and internalised stigma, while fear of discrimination can delay diagnosis, reduce healthcare engagement and worsen disability and social exclusion.

Leprosy is frequently complicated by immune-mediated inflammatory episodes known as leprosy reactions which may occur before, during or after completion of anti-microbial treatment (1). Leprosy reactions are the most important risk factor for nerve damage and disability(11,12). Erythema nodosum leprosum (ENL) or leprosy Type 2 reaction, is a severe multisystem inflammatory complication affecting individuals with borderline lepromatous leprosy and lepromatous leprosy (12,13). ENL is characterised by episodes of painful erythematous cutaneous nodules accompanied by fever, peripheral oedema, arthritis, neuritis, iridocyclitis and orchitis (14). These episodes may be acute, recurrent or chronic, often requiring prolonged treatment with immunosuppressant drugs and repeated healthcare appointments (11–13). ENL predominantly affects male adults of working age (14,15) and can result in substantial disability, reduced health-related quality of life (HRQoL) and significant economic burden (16–18).

Measurement of HRQoL has gained increasing importance in neglected tropical diseases, including leprosy, where long-term morbidity, disability and stigma substantially affect well-being and social participation (19,20). Several instruments have been used to evaluate HRQoL in leprosy, including World Health Organization Quality of Life questionnaire (WHOQoL), the 36-Item Short Form Survey (SF-36) and the Dermatology Life Quality Index (DLQI)(6,21–24).

The DLQI is a widely used dermatology-specific questionnaire designed to assess the impact of skin disease on the HRQoL of individuals (16 years and older) in the preceding week (25). It consists of ten items covering symptoms and feelings, daily activities, leisure, work and school, personal relationships, and treatment burden. Scores range from 0 to 30, with higher scores indicating greater impact (25). The DLQI is extensively used in dermatology practice and research (26), including in the United Kingdom where it is incorporated into clinical guidelines to guide treatment decisions for conditions such as psoriasis (27). The DLQI is practical, brief and easy to administer in clinical settings.

The DLQI was not developed specifically for leprosy but has been used in studies of people affected by the disease, with variable findings depending on the type of leprosy, disability status and socioeconomic context (28–30). Cross-sectional studies using DLQI have demonstrated poorer HRQoL amongst individuals experiencing leprosy reactions, including ENL, compared with those without reactions (28,31). However, ENL is a multisystem inflammatory condition with extra-cutaneous manifestations (14) and the DLQI may not fully capture the broader physical and psychological burden of ENL(32). Generic HRQoL instruments, such as the SF-36, have shown substantial impairment in both physical and mental health domains among individuals with ENL (16,17,33). The SF-36 has been validated in people affected by leprosy(34).

(32) Determining whether a short dermatology-specific instrument can adequately reflect broader HRQoL is important for selecting outcome measures in ENL research and clinical practice. Evidence examining the relationship between the DLQI and SF-36 in people affected by leprosy remains limited. Only one study with 104 participants with leprosy explored the correlation between DLQI and SF-36, and reported a weak association, suggesting the DLQI may capture only part of the overall health impact of leprosy on HRQoL (28). The Methotrexate and Prednisolone study in ENL (MaPs in ENL), was a multicentre international double-blind randomised clinical trial conducted in Ethiopia, India, Indonesia, and Nepal with participants with severe ENL (35). In the trial, the DLQI and SF-36 were completed at four predefined timepoints to assess HRQoL. Higher DLQI scores indicate greater impairment in HRQoL, whereas lower SF-36 scores indicate worse health status. Differences in HRQoL between intervention arms and changes over time were evaluated as secondary outcomes of the trial and have been reported previously(35).

The aim of this study was to examine the relationship between the dermatology-specific DLQI and the generic SF-36 using the large, well characterised longitudinal data from MaPs in ENL to determine the validity of the DLQI in measuring HRQoL in individuals affected by ENL.

## METHODS

### Study design and setting

This study is a post-hoc analysis of prospectively collected data from MaPs in ENL (35). For the present analysis, participants were considered as a longitudinal cohort to compare HRQoL measures.

Participants were recruited from five specialist leprosy referral centres in four countries: the ALERT Comprehensive Specialized Hospital, Addis Ababa, Ethiopia; The Leprosy Mission Trust India Barabanki Hospital, Barabanki and Bombay Leprosy Project, Mumbai, India; Dr. Soetomo Hospital, Surabaya, Indonesia and The Leprosy Mission Nepal Anandaban Hospital, Kathmandu, Nepal.

### Participants

Participants were individuals with severe ENL (ENLIST ENL Severity Scale score of nine or more)(36) who were screened for enrolment in the MaPs trial and met the trial eligibility criteria (35). Participants were randomised to receive weekly oral methotrexate and a 20-week course of daily prednisolone or placebo and prednisolone. HRQoL was assessed at predefined timepoints (enrolment, 24, 48 and 60 weeks)(35). Data from all participants were analysed regardless of intervention allocation, as this analysis aimed to assess the performance of DLQI rather than differences between intervention groups. Eligibility was assessed according to the predefined inclusion and exclusion criteria described in the published trial protocol (35).

For descriptive and subgroup analyses, ENL episodes were classified according to their clinical course as acute or recurrent or chronic, based on the definitions specified in the trial protocol (35). For this analysis, participants recruited at the Ethiopian centre were excluded because a different version of the SF-36 was used. To ensure comparability of the SF-36 across centres, the present analysis was restricted to participants recruited in India, Indonesia and Nepal.

### Health-related quality of life assessments

HRQoL was assessed using : the DLQI (25,37) and SF-36 (38). Validated translations of both questionnaires in the first language of the participant were administered at enrolment and follow-up visits at weeks 24, 48, and 60(37,39–43).

DLQI scores greater than 10 indicate a very large or extremely large impact on HRQoL (25). Table 1 presents the interpretation of the DLQI scores.

**Table 1:**
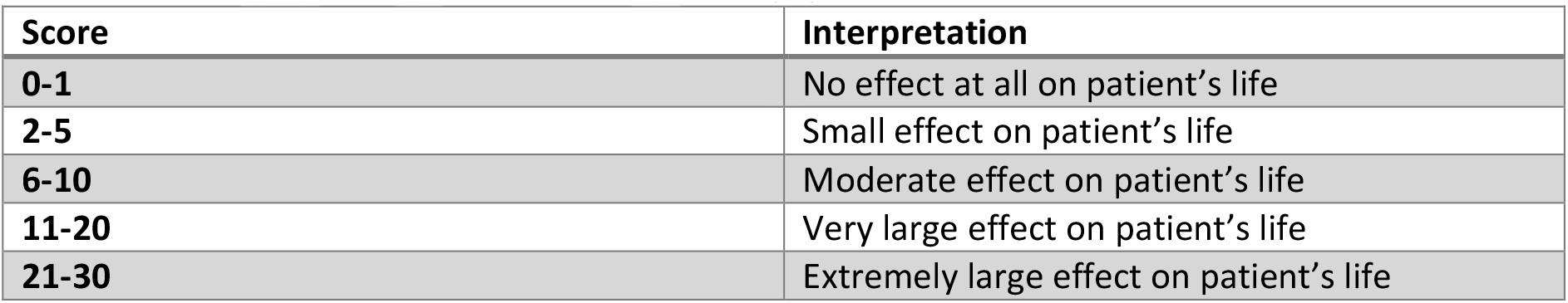
Dermatology Life Quality Index scores interpretation(25)

| Score | Interpretation |
| --- | --- |
| 0-1 | No effect at all on patient's life |
| 2-5 | Small effect on patient's life |
| 6-10 | Moderate effect on patient's life |
| 11-20 | Very large effect on patient's life |
| 21-30 | Extremely large effect on patient's life |

The SF-36 evaluates eight domains of health, including physical functioning, bodily pain, general health, vitality, social functioning and mental health. Domain scores range from 0 to 100, with higher scores representing better perceived health status (38). Two standardised summary measures are derived from the SF-36: the Physical Component Summary (PCS) and the Mental Component Summary (MCS). These indices aggregate the eight SF-36 domains into two overarching dimensions representing physical and mental health, allowing a more parsimonious assessment of overall HRQoL and facilitation of comparisons across studies and populations (38).

### Data collection

Data were entered prospectively into a trial-specific database using Research Electronic Data Capture (REDCap) (Vanderbilt University, Nashville, TN, USA), a secure data management platform compliant with Good Clinical Practice standards. Data quality procedures included range checks, validation rules and routine data cleaning prior to analysis. Analyses were conducted using available data for each timepoint without imputation.

### Statistical analysis

Descriptive statistics of continuous variables were calculated using means and standard deviations (SD) or medians and interquartile rages (IQR). DLQI scores were positively skewed and are therefore presented as medians with IQR, whereas SF-36 components scores are presented as means with SD. Associations between DLQI and SF-36 scores were examined using Pearson correlation coefficients, with Spearman rank correlations used as a robust alternative for the skewed distribution of DLQI scores. Comparisons between ENL subtypes (acute vs recurrent or chronic) were performed using Wilcoxon rank-sum tests for continuous variables. At enrolment, the relationship between DLQI and SF-36 PCS and MCS scores was examined using multivariable linear regression models adjusted for age, sex and recruiting centre with robust standard errors to account for potential non-constant variance in the residual (heteroscedasticity).Longitudinal associations during follow-up were examined using linear mixed-effects models with a random intercept for participant to account for repeated measurements within individuals. Time was modelled as a categorical fixed effect (week 24, week 48 and week 60) and an interaction term between DLQI and timepoint was included to assess whether the association between DLQI and SF-36 varied over time. These models were adjusted for age, sex, recruiting centre and the enrolment value of the respective SF-36 summary score to account for initial differences and regression to the mean. Marginal effects were estimated to quantify the association between DLQI and SF-36 scores at each follow-up timepoint. All tests were two-sided and p < 0.05 was considered statistically significant. All statistical analyses were performed using STATA (version 18 StataCorp, College Station, TX, USA).

## RESULTS

A total of 129 participants were included in the HRQoL analysis at enrolment. During follow-up, 383 HRQoL assessments were available at four timepoints: at enrolment (n=129), week 24 (n=99), week 48 (n=77) and week 60 (n=78). Table 2 summarises the characteristics of participants who were included in this study.

**Table 2:** Characteristics of participants at enrolment.

| Characteristics | At enrolment (n=129) |
| --- | --- |
| Mean Age, years | 33.8 (SD 10.4) |
| Sex (%) Male | 97 (75.2) |
| ENL classification, n (%) |  |
| Acute | 39 (30.2) |
| Recurrent or Chronic | 90 (69.8) |
| Health Related Quality of Life instruments |  |
| Median DLQI score | 19 (IQR 15–21) |
| DLQI >10, n (%) | 109 (84.5) |
| Mean SF-36 Physical Component Summary | 30.3 (SD 7.3) |
| Mean SF-36 Mental Component Summary | 33.3 (SD 8.4) |

**Abbreviations:** SD, standard deviation; IQR, interquartile range; DLQI, Dermatology Life Quality Index; SF-36, 36-Item Short Form Health Survey.

DLQI scores were positively skewed. At enrolment, DLQI scores indicated a very large effect on participants life with a median of 19 (IQR 15-21), consistent with severe ENL prior to treatment. DLQI scores improved markedly at follow-up with medians scores decreasing to 3 (IQR 1-10) at week 24, 2 (IQR 0-7) at week 48 and 1.5 (IQR 0-5) at week 60. The distribution of DLQI median scores over time is shown in Figure 1.

**Figure 1:**
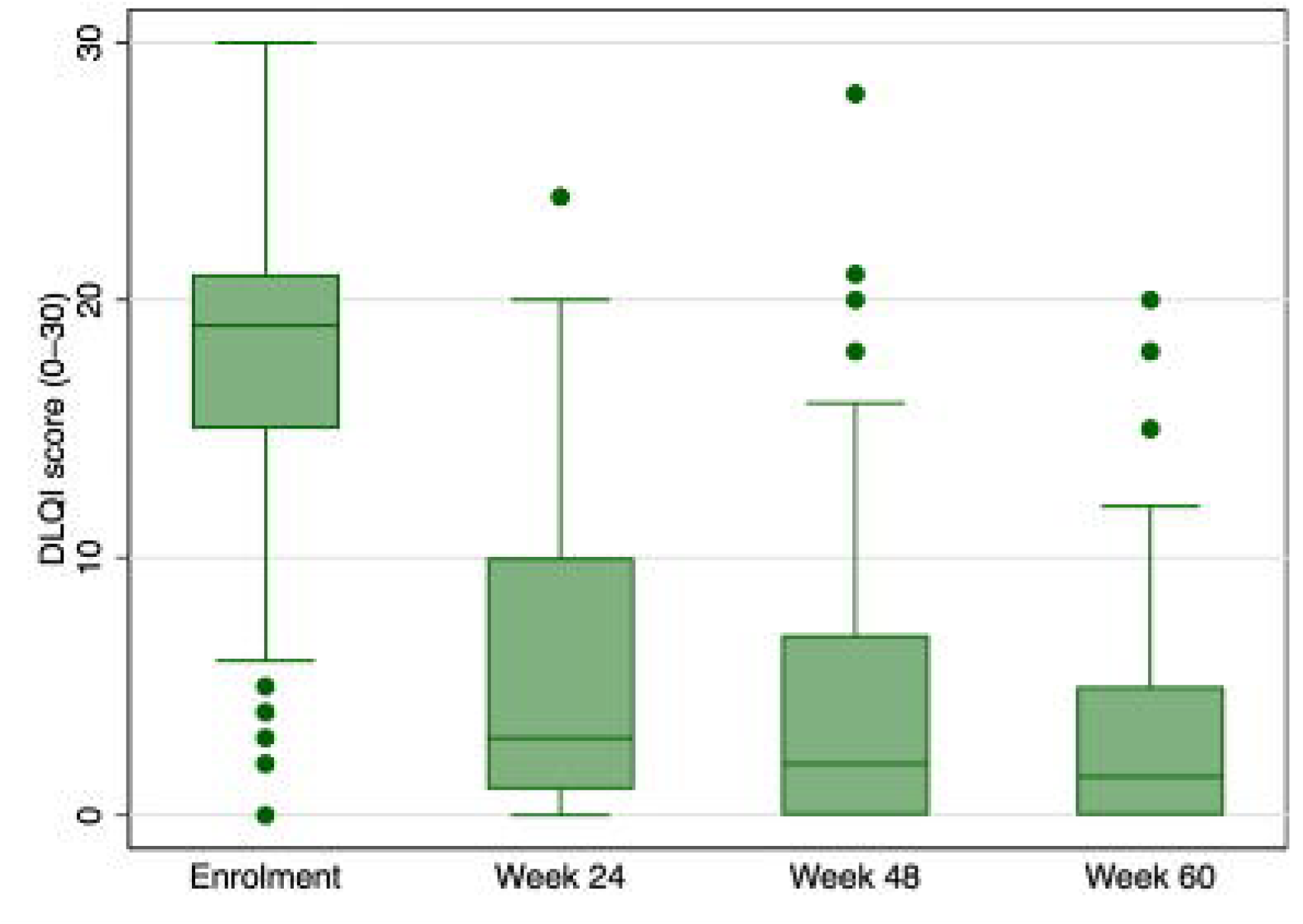
Box plots of Dermatology Life Quality Index scores at enrolment and follow-up visits, showing median values and score distribution in participants in the Methotrexate and Prednisolone study in erythema nodosum leprosum.

Mean SF-36 component scores increased substantially after enrolment, indicating an improvement in HRQoL. At enrolment mean PCS was 30.3 (SD 7.3) and the mean MCS was 33.3 (SD 8.4). At week 24 PCS increased to 46.5 (SD 9.1) and MCS 47.8 (SD10.7). At week 48, mean PCS was 48.6 (SD 8.6) and MCS 50.4 (SD 8.4), and by week 60 PCS 49.3 (SD 8.1), MCS 50.9 (SD 9.9). Overall, both physical and mental health status improved from enrolment. Table 3 summarises DLQI and SF-36 scores across all timepoints.

**Table 3:** Dermatology Life Quality Index and SF-36 scores across follow up timepoints.

| Timepoint | DLQI median (IQR) | PCS mean (SD) | MCS mean (SD) |
| --- | --- | --- | --- |
| At enrolment | 19 (15–21) | 30.26 (7.30) | 33.31 (8.39) |
| Week 24 | 3 (1–10) | 46.46 (9.09) | 47.78 (10.70) |
| Week 48 | 2 (0–7) | 48.63 (8.60) | 50.42 (8.42) |
| Week 60 | 1.5 (0–5) | 49.34 (8.08) | 50.95 (9.85) |

**Notes:** DLQI presented as median (IQR) due to skewness; PCS/MCS as mean (SD).

### Correlation between DLQI and SF-36 summary scores

DLQI scores were strongly and consistently associated with SF-36 summary measures of HRQoL. Higher DLQI scores were associated with lower SF-36 PCS and MCS scores. Spearman rank correlation coefficients demonstrated moderate to strong negative correlations between DLQI and both PCS and MCS. These associations were consistent across all follow-up timepoints, indicating that worse dermatology-specific HRQoL was associated with worse overall physical and mental health status. Similar patterns were observed across individual SF-36 domains, with DLQI showing negative correlations with domains related to physical functioning, bodily pain, vitality, social functioning and mental health.

Across all timepoints pooled (n=383), DLQI was strongly inversely correlated with both SF-36 summary measures. The Pearson correlation between DLQI and PCS was r = −0.848 (p<0.001), and between DLQI and MCS was r = −0.806 (p<0.001). Spearman correlations were similar (ρ = −0.839 for DLQI–PCS and ρ = −0.775 for DLQI–MCS; both p<0.001), supporting a robust monotonic association despite the skewed distribution of DLQI at follow-up. When examined by timepoint, negative correlations were observed at all visits (Table 4). At enrolment, correlations were moderate (DLQI–PCS r=−0.569; DLQI–MCS r=−0.533; both p<0.001). From week 24 onwards, correlations strengthened and remained high (DLQI–PCS r=−0.756 to −0.755; DLQI–MCS r=−0.721 to −0.705; all p<0.001).

**Table 4:**
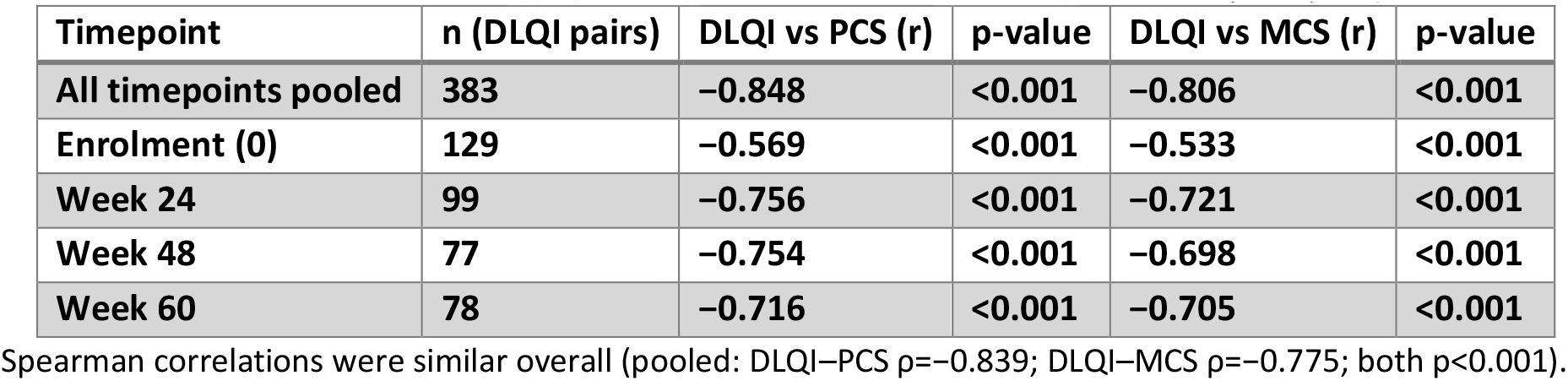
Correlation between Dermatology Life Quality Index and summary scores (pooled and by timepoint)

| Timepoint | n (DLQI pairs) | DLQI vs PCS (r) | p-value | DLQI vs MCS (r) | p-value |
| --- | --- | --- | --- | --- | --- |
| All timepoints pooled | 383 | -0.848 | <0.001 | -0.806 | <0.001 |
| Enrolment (0) | 129 | -0.569 | <0.001 | -0.533 | <0.001 |
| Week 24 | 99 | -0.756 | <0.001 | -0.721 | <0.001 |
| Week 48 | 77 | -0.754 | <0.001 | -0.698 | <0.001 |
| Week 60 | 78 | -0.716 | <0.001 | -0.705 | <0.001 |

Spearman correlations were similar overall (pooled: DLQI–PCS $\rho = -0.839$ ; DLQI–MCS $\rho = -0.775$ ; both $p < 0.001$ ).

Visual inspection using a locally weighted scatterplot smoothing (LOWESS) curves demonstrated an approximately linear relationship, supporting the use of linear models for analysis. Figure 2 illustrates the inverse relationship between DLQI and PCS across all study timepoints. Figure 3 illustrates the inverse relationship between DLQI and MCS across all study timepoints.

**Figure 2:**
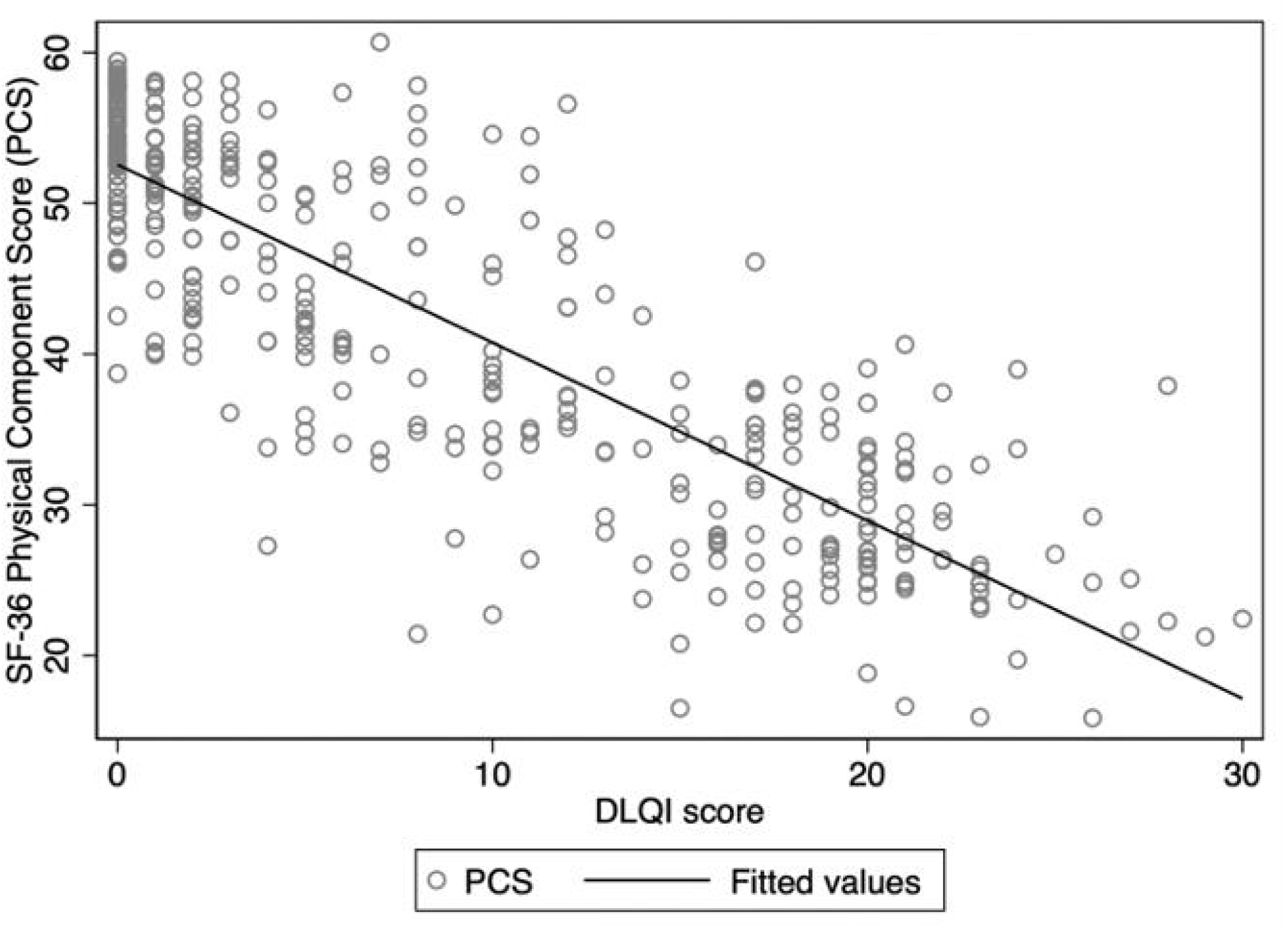
Scatter plot showing the association between Dermatology Life Quality Index score and SF-36 Physical Component Summary score in participants in the Methotrexate and Prednisolone study in erythema nodosum leprosum.

**Figure 3:**
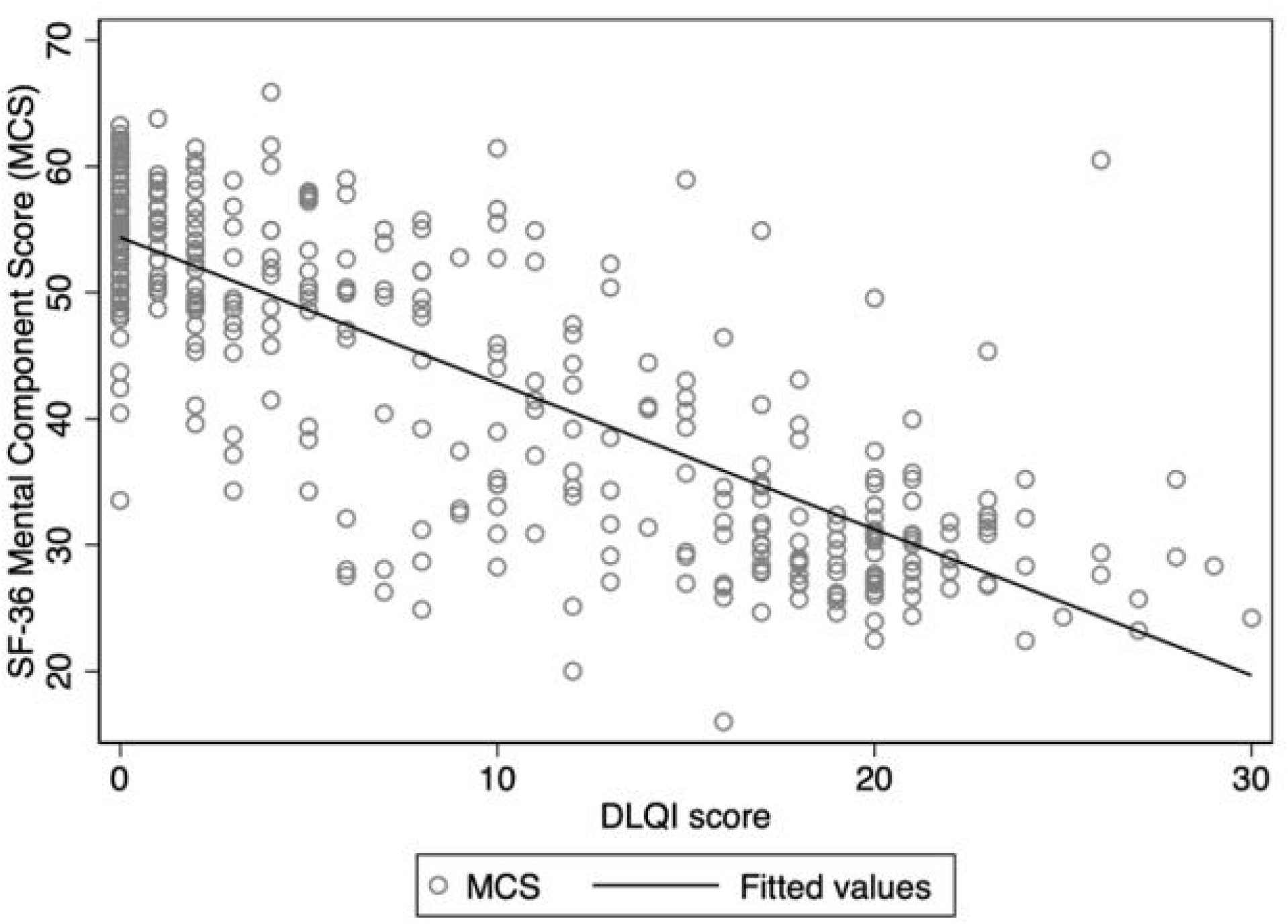
Scatter plot showing the association between Dermatology Life Quality Index score and SF-36 Mental Component Summary score across all study timepoints in participants in the Methotrexate and Prednisolone study in erythema nodosum leprosum.

### Correlation between DLQI and SF-36 domains

The DLQI was consistently inversely correlated with all eight SF-36 domains at each timepoint (Table 5). The strongest associations were generally observed for domains reflecting physical limitation and symptom burden (e.g., bodily pain, vitality, physical functioning) and for mental health at follow-up. Correlations with general health were weaker than for other domains, particularly at week 60 (DLQI–GH r=−0.343; p=0.002).

**Table 5:** SF-36 domain correlations with DLQI by timepoint (Pearson correlation coefficient)

| Domain | Enrolment<br>(Pearson<br>correlation<br>coefficient) | Week 24<br>(Pearson<br>correlation<br>coefficient) | Week 48<br>(Pearson<br>correlation<br>coefficient) | Week 60<br>(Pearson<br>correlation<br>coefficient) |
| --- | --- | --- | --- | --- |
| Physical Functioning (PF) | -0.594 | -0.730 | -0.712 | -0.695 |
| Role Physical (RP) | -0.550 | -0.745 | -0.596 | -0.759 |
| Role Emotional (RE) | -0.451 | -0.723 | -0.492 | -0.757 |
| **Vitality (VT)** | -0.568 | -0.687 | -0.806 | -0.714 |
| **Mental Health (MH)** | -0.600 | -0.673 | -0.787 | -0.716 |
| **Social Functioning (SF)** | -0.315 | -0.676 | -0.657 | -0.628 |
| **Bodily Pain (BP)** | -0.494 | -0.714 | -0.768 | -0.700 |
| **General Health (GH)** | -0.412 | -0.432 | -0.462 | -0.343 |

### Comparison by ENL clinical course (acute vs recurrent or chronic)

ENL classification was available for 383 assessments (acute n=110, recurrent/chronic n=273). At enrolment, DLQI scores were higher amongst participants with acute ENL compared with recurrent or chronic ENL (Wilcoxon rank-sum p=0.013), indicating worse dermatology-specific HRQoL at presentation. Acute ENL was also associated with lower PCS at enrolment (p=0.001), whilst there was no evidence of a difference between acute and chronic or recurrent ENL cases in MCS at enrolment(p=0.115). At follow-up (weeks 24, 48, and 60), there was no evidence of differences in DLQI by ENL classification (all p>0.30). PCS did not differ by ENL classification at weeks 24 or 48; however, at week 60, PCS was significantly higher in those with acute ENL compared to those with recurrent or chronic ENL (p=0.016). No differences in MCS were observed at any timepoint (all p≥0.21).

### Association between DLQI and SF-36 summary scores

At enrolment, higher DLQI scores were associated with worse HRQoL. In adjusted linear regression models, each 1-point increase in DLQI was associated with a 0.66-point reduction in PCS (95% CI −0.884 to −0.436; p<0.001) and a 0.51-point reduction in MCS (95% CI −0.755 to −0.265; p<0.001). In longitudinal mixed-effects models accounting for repeated measurements within participants and adjusted for enrolment SF-36 scores, higher DLQI scores remained strongly associated with lower PCS and MCS during follow-up.

For PCS, each 1-point increase in DLQI was associated with reductions of 1.15 points at week 24, 1.04 points at week 48, and 1.17 points at week 60 (all p<0.001). There was no evidence that the strength of this association varied over time (interaction p=0.605). Similarly, for MCS, each 1-point increase in DLQI was associated with reductions of 1.28 points at week 24, 0.99 points at week 48, and 1.22 points at week 60 (all p<0.001), with no evidence of interaction between DLQI and timepoint (p=0.225).

## DISCUSSION

This post-hoc analysis of the MaPs in ENL dataset demonstrates a strong association between the dermatology-specific DLQI and generic SF-36 measures of HRQoL. Higher DLQI scores were consistently associated with lower SF-36 PCS and MCS scores at enrolment and during follow-up, indicating that worsening dermatology-related HRQoL reflects poorer overall health status in people with ENL.

The negative correlation observed between DLQI and the SF-36 summary components is expected given the inverse scoring system of the scales: higher DLQI scores indicate worse quality of life, whilst higher SF-36 scores indicate better health status. When the eight individual SF-36 domains were examined separately, the strength of correlation with DLQI was lower. This is not unexpected, as the domains of the SF-36 assess a broad range of aspects of health including physical functioning, emotional well-being, social functioning and vitality, whereas DLQI is specifically designed to capture the impact of skin disease on daily life (38).

The association between DLQI and the SF-36 summary components persisted after adjustment for potential confounders and enrolment HRQoL and remained stable across follow-up timepoints. This indicates that dermatology-specific impairment is consistently associated with overall physical and mental health status, independent of initial differences and repeated measurements within individuals. These findings support the validity of using DLQI as a proxy indicator of broader health impact in individuals affected by ENL.

Participants with acute ENL had significantly worse DLQI and physical health component scores at enrolment compared with those with recurrent or chronic ENL. These differences did not persist during follow-up, suggesting that the acute inflammatory phase of ENL may have the greatest impact on dermatological symptoms and physical functioning. An alternative explanation is adaptation to chronic disease(44,45), whereby individuals with longstanding ENL may report better HRQoL over time despite ongoing symptoms, a phenomenon described in other chronic dermatological conditions such as psoriasis(46).

The painful clinical manifestations of ENL, including painful skin nodules, fever, arthritis, neuritis, malaise, oedema and lymphadenitis, are likely to contribute substantially to impaired physical well-being and limitations in daily functioning (16,17,28,33). Previous cross-sectional studies have demonstrated that ENL is associated with significant impairment of HRQoL across multiple domains (16,17,33). In Bangladesh, significantly worse HRQoL was reported across all eight domains of the SF-36 in individuals with ENL compared with those with multibacillary leprosy without reactions (16). Similarly, a study conducted in Brazil found that patients with acute ENL had markedly reduced physical and mental health scores particularly in domains related to bodily pain and physical functioning (33).

A cross-sectional study conducted in Malaysia with 153 individuals with multibacillary leprosy reported significantly higher DLQI scores amongst individuals with ENL compared with individuals without reaction (31). The domains most affected included symptoms and feelings as well as daily activities, highlighting the significant burden imposed by ENL.

The correlation between DLQI and SF-36 observed in the present study suggests that dermatology specific impairment reflects broader aspects of health status in individuals with ENL. However, a previous cross-sectional study from Brazil evaluating the relationship between these instruments in a heterogeneous leprosy population (including individuals with paucibacillary and multibacillary disease, at different stages of treatment, with varying degrees of disability and presence or absence of reactions) reported weaker correlations between DLQI and SF-36 domains (28). The severe, painful inflammatory nature of ENL in general, and in our cohort in particular may explain (14) the stronger association between dermatology-specific and generic measures of HRQoL in ENL compared to other clinical presentations of leprosy. The persistence of this association across all timepoints further supports the construct validity of DLQI as a measure of HRQoL in ENL.

These findings highlight the considerable reduction in HRQoL associated with ENL and support the use of the DLQI to assess this. The study of Guimenes Albuquerque et al. may have showed weaker correlations between the DLQI and SF-36 because it had a heterogeneous cohort of 104 individuals with and without leprosy reactions. Only 14.4% had ENL and 9.6% were described as having a mixed pattern of leprosy reactions (Type 1 and ENL)(28).

The DLQI is available in more than 140 translations.(37) It is simple to administer, easy to score and straightforward to interpret, which makes it particularly suitable for routine clinical practice. Although the SF-36 provides a broader assessment of physical and mental health, the strong correlation observed between the two instruments suggests that the DLQI can successfully capture wider aspects of HRQoL of individuals with ENL.

This study has several strengths. It uses data from MaPs in ENL, a multicentre prospective clinical trial with standardised data collection and repeated HRQoL assessments. The availability of paired DLQI and SF-36 scores enabled direct comparison between dermatology-specific and generic measures of HRQoL in individuals with ENL. However, this was a secondary analysis, and the study was not designed to evaluate the relationship between HRQoL instruments. The number of completed HRQoL assessments decreased after enrolment, with paired assessment declining from 129 at enrolment to 78 at week 60, which may have reduced statistical power. In addition, participants were recruited within a clinical trial and may not fully represent individuals with ENL managed in routine clinical settings or fully reflect the HRQoL of those participants who withdrew from the trial.

## CONCLUSION

The DLQI is a practical tool to assess the HRQoL and has a strong correlation with generic HRQoL measures. The DLQI is a useful patient reported outcome measure for evaluating HRQoL and its association with treatment in future ENL clinical studies and routine clinical practice.

## ACKNOWLEDGEMENTS

We would like to thank the people with lived experience of leprosy who participated in this study. We thank the clinical and research staff at all participating centres for their dedication to recruitment, follow-up, and patient care. We acknowledge the contribution of the ENLIST Group, including Marivic Balagon, C. Ruth Butlin, Milton O. Moraes (in memoriam), Jose da Costa Nery, and Anna Sales, for their continued commitment to advancing research in ENL and improving outcomes for affected communities.

## AUTHOR CONTRIBUTION

Conceptualisation: Barbara de Barros, Diana N. J. Lockwood, Stephen L. Walker and the Erythema Nodosum Leprosum International Study Group (ENLIST). Methodology: Barbara de Barros, Bernd Genser, Peter Nicholls, Diana N. J. Lockwood and Stephen L. Walker. Formal Analysis: Barbara de Barros, Stephen L. Walker and Bernd Genser. Investigation: Farha Sultana, Anju Wakade, Bhagyashree Bhame, Bishwanath Acharya, Abdulnaser Hamza, Alemtsehay Getachew, Medhi Denisa Alinda, M. Yulianto Listiawan, Shimelis N Doni, Deanna A. Hagge, Indra Napit, Mahesh Shah, Vivek V. Pai, Neeta Maximus, Joydeepa Darlong. Data Curation: Barbara de Barros, Bernd Genser, Peter Nicholls. Project Administration: Barbara de Barros. Supervision: Diana N. J. Lockwood and Stephen L. Walker. Writing – Original Draft Preparation: Barbara de Barros. Writing – Review & Editing: All authors. Funding Acquisition: Diana N. J. Lockwood, Stephen L. Walker

## DATA AVAILABILITY

The data underlying this study will be deposited in the London School of Hygiene & Tropical Medicine (LSHTM) Data Repository. The dataset will be available following an embargo period of one year from the date of publication. After this period, data may be accessed upon reasonable request and in accordance with LSHTM data sharing policies.

